# Evaluation of SARS-CoV-2 IgG antibody response in PCR positive patients: comparison of nine tests in relation with clinical data

**DOI:** 10.1101/2020.07.15.20149617

**Authors:** Paul Naaber, Kaidi Hunt, Jaana Pesukova, Liis Haljasmägi, Pauliina Rumm, Pärt Peterson, Jelena Hololejenko, Irina Eero, Piia Jõgi, Karolin Toompere, Epp Sepp

**Author notes:** Corresponding author; SYNLAB Estonia, Veerenni 53a, 11313 Tallinn, Estonia.

## Abstract

**Background:** High number of SARS-CoV-2 antibody tests are available in different formats, detect different types of antibodies, and use different target proteins. Sensitivity of these tests varies and could be also related to clinical symptoms and testing time.

**Methods:** Serum samples from 97 COVID-19 patients and 100 controls were tested with 9 antibody tests (SNIBE, Epitope, Euroimmun, Roche, Abbott, DiaSorin, Biosensor, LIPS N, and LIPS S-RBD). The results were analyzed in context of clinical data.

**Findings:** Positivity rate was of tests was following: N-LIPS test (91.8% cases), Epitope (85.6%), Abbott and in-house LIPS S-RBD (both 84.5%), Roche (83.5%), Euroimmun (82.5%), DiaSorin (81.4%), SNIBE (70.1%), and Biosensor (64.9%). Agreement between tests varied (71-95%). Correlation between patient symptoms score and antibody value was test-dependent: varied from strongest in LIPS N (ρ=0.41; p<0.001) to nonsignificant (LIPS S-RBD). Testing time from symptoms influenced sensitivity in some tests more than another’s.

**Interpretation:** Sensitivity of tests varied highly and combination of different tests may improve it. Relation of results to symptoms and testing time was test-dependent. Thus, some antibody tests seems to be more sensitive to detect antibodies early and in asymptomatic patients than others.

**Funding:** Study was funded by SYNLAB Estonia and Estonian Research Council grant PRG377.

**Research in context:** *Evidence before this study:* High variation in COVID-19 antibody tests sensitivity has been described. Relation between antibody response and clinical cause has been found in some studies but not in others. It’s known that antibody response is time dependent, however seroconversion medians varied in different studies.

*Added value of this study:* We confirmed that SARS-CoV-2 antibody response depends on clinical symptoms and time of testing, but we also found that this relation is dependent on test setup and viral antigens used in the tests. This may explain contradictory results of previous studies.

*Implications of all the available evidence:* Our study has practical implications showing that not all antibody tests work uniformly well in symptomatic and asymptomatic cases and in different time periods from disease onset. This should be taken into consideration in clinical practice for diagnosing COVID-19 and in epidemiological studies evaluating the seroprevalence especially in asymptomatic population.

## Introduction

More than 300 tests are available to detect SARS-CoV-2 antibodies (1). These tests are produced in different formats and detect different types of antibodies including IgG, IgM or IgA subtypes or total immunoglobulin. In addition, the target proteins used to detect antibodies vary between the tests, but they most often include nucleocapsid (N), spike1 (S1), spike2 (S2), receptor binding domain of the spike (S-RBD) protein, or their combinations. Although the producers usually have reported high sensitivity and specificity for their tests, variable clinical sensitivity has been reported by independent studies (2-5). However, minimal data is available on their sensitivity and specificity in respect to their differences in target proteins.

Majority of clinical studies and validations of commercial tests have studied the patients with severe disease and only few have investigated the antibody responses in pauci-symptomatic or asymptomatic persons (6,7). Several studies have shown stronger antibody response in patients with severe disease as compared with mildly symptomatic ones and also higher rate of absence of seroconversion in asymptomatic patients. However, other studies have failed to find any correlation between clinical course and immune response (3,6). Thus, the reported sensitivity of individual tests may depend on a selected group of patients used for test evaluation.

It is known that the sensitivity of antibody test depends on a sampling time. Different studies have reported variable time of appearance of antiviral IgG antibodies but in most publications the median seroconversion time has been between 6 and 14 days from symptoms onset. Although several studies have shown high IgG at least for seven weeks, rapid decline of IgG in convalescence phase has been reported in asymptomatic COVID-19 patients (2,3,6). Thus, the optimal time for IgG detection (with the highest sensitivity rate) may also depend on clinical course of COVID-19 and is not clearly defined yet.

## Aim

Aim of our study was to compare the performance characteristics of 7 commercial and 2 in-house IgG/total Ab tests, which analyze the reactivity to several target proteins, and to correlate the results with the patients clinical data (with different symptoms score, age, and time from disease onset).

## Material and methods

### Patients and samples

Serum samples from 97 persons with COVID-19 were collected between April 28 and May 07 2020 in Saaremaa, Estonia (the island with highest rate of SARS-CoV-2 detected cases in Estonia, 166.4 cases per 10 000 inhabitants (8). The inclusion criterion was confirmed by positive SARS-CoV-2 RT-PCR test. All patients signed informed consent form and filled questionnaire about clinical data. Patients’ data is shown in Supplementary table 1.

For testing the specificity 100 anonymous serum samples were collected in 2019 before COVID-19 pandemic and stored in SYNLAB Estonia.

### Tests

All samples were stored at – 30° C and had one freezing/thawing cycle before testing. Five laboratory tests for IgG (SNIBE, Euroimmun, Abbott, Epitope, DiaSorin), one laboratory total Ab (Roche) test, one rapid IgG test (SD Biosensor) and 2 in-house IgG tests (LIPS N and LIPS S-RDB) were included into comparison. Different protein markers have been used in these tests (S1, S2, S-RBD, N or their combinations). Detailed information about the tests is summarized in Supplementary table 2. Commercial tests were performed and interpreted according to manufacturer instruction, in-house LIPS as described previously (9).

### Statistics

The following analysis was made: correlation by Spearman’s test, comparison of qualitative data (positivity rate) by Fisher exact test, groups’ comparison by Kruskal-Wallis test, pairwise comparison by Conover-Iman test with Holm-Bonferron correction using Past 4.03 and Stata 14.2 software.

## Results

### Comparison of specificity of tests

Testing of 100 pre COVID-19 sera by Roche, Abbott and Biosensor gave no false positive results. DiaSorin gave 1 and SNIBE 2 false positive results; Euroimmun 2 and Epitope 2 false positive and accordingly 2 and 4 borderline results using the producers’ interpretation criteria. Both LIPS tests gave 2 false positive results using arbitrary cut-offs. In total, 15 (15%) control samples gave false positive result by any test. In most cases only one test was positive per sample but 2 samples showed positive/borderline result by two (Epitope and Euroimmun) or three (SNIBE, Epitope, S-RBD LIPS) tests. Distribution of quantitative values of controls is presented in Supplementary figure 1.

### Comparison of sensitivity by different tests in COVID-19 patients

53 (55%) out of 97 COVID-19 patients samples were positive and 2 (2%) negative by all applied tests for IgG or total SARS-CoV-2 antibodies. Remaining 42 samples (43%) were positive by some of the tests. The highest positivity rate was found in in-house N-LIPS test (91.8% cases), followed by Epitope (85.6%), Abbott and in-house LIPS S-RBD (both 84.5%), Roche (83.5%), Euroimmun (82.5%), DiaSorin (81.4%), SNIBE (70.1%) and Biosensor rapid test (64.9%).

The agreement between the tests (using qualitative interpretations) ranged between 95% (Euroimmun and DiaSorin) and 71% (LIPS N and Biosensor). Correlation between quantitative results was significant in all tests combinations (p<0.001) except between in-house N and S-RBD LIPS that gave non-significant result. The strongest correlation was found between Euroimmun and DiaSorin tests (ρ=0.95). Agreement between qualitative results and correlation between quantitative values is shown in Table 1 and Supplementary figure 2. Distribution of COVID-19 cases tests values in comparison with control samples is presented in Supplementary figure 1.

**Table 1.** Agreement between tests in COVID-19 patients (n=97)

| Tests | Agreement between qualitative results, % |  |  |  |  |  |  |  |
| --- | --- | --- | --- | --- | --- | --- | --- | --- |
| | Correlation between quantitative values, $\rho$ ($p < 0.001$ in all significant cases) | | | | | | | |
|  | Epitope | Euroimmun | Roche | Abbott | DiaSorin | LIPS RBD | LIPS N | Biosensor |
| SNIBE | 85 | 77 | 85 | 86 | 80 | 84 | 76 | 85 |
|  | 0.91 | 0.64 | 0.68 | 0.84 | 0.70 | 0.6 | 0.56 | NA |
| Epitope |  | 82 | 90 | 93 | 84 | 91 | 86 | 77 |
|  |  | 0.56 | 0.59 | 0.80 | 0.65 | 0.53 | 0.46 | NA |
| Euroimmun |  |  | 87 | 81 | 95 | 88 | 85 | 74 |
|  |  |  | 0.66 | 0.65 | 0.95 | 0.73 | 0.50 | NA |
| Roche |  |  |  | 89 | 88 | 91 | 88 | 79 |
|  |  |  |  | 0.78 | 0.71 | 0.67 | 0.51 | NA |
| Abbott |  |  |  |  | 82 | 90 | 87 | 78 |
|  |  |  |  |  | 0.71 | 0.59 | 0.51 | NA |
| DiaSorin |  |  |  |  |  | 87 | 84 | 77 |
|  |  |  |  |  |  | 0.78 | 0.53 | NA |
| LIPS S-RBD |  |  |  |  |  |  | 80 | 76 |
|  |  |  |  |  |  |  | NS | NA |
| LIPS N |  |  |  |  |  |  |  | 71 |
|  |  |  |  |  |  |  |  | NA |

NS= not significant correlation; NA – not applicable

### Relations between antibody detection and COVID-19 patients’ symptoms

Correlating the patients’ symptom scores and quantitative values of the tests, we found significant positive correlation in all cases except with LIPS S-RBD. The strongest correlation was found in LIPS N (ρ=0.41; p<0.001), followed by Roche (ρ=0.39; p<0.001), Abbott and SNIBE (both ρ=0.32; p=0.001), DiaSorin (ρ=0.31; p=0.002), Epitope (ρ=0.30; p=0.003) and Euroimmun (ρ=0.29; p=0.004). The differences in quantitative test values and qualitative results among patient groups according to the number of symptoms are presented in table 2.

**Table 2.** Antibody detection by tests in COVID-19 patients (n=97) with different symptoms score

| Tests | Quantitative test value median (25%; 75% percentiles)<br>% of positive tests in subgroup |  |  |
| --- | --- | --- | --- |
|  | Asymptomatic<br>n=20 | Symptoms score 1-6<br>n=43 | Symptoms score 7-14<br>n=34 |
| SNIBE | 0.79 (0.16; 12.6)<sup>1</sup><br>40<sup>#,□</sup> | 2.39 (0.97; 12.11)<br>74<sup>#</sup> | 9.05 (2.00; 21.7)<sup>1</sup><br>82<sup>□</sup> |
| Epitope | 0.47 (0.26; 0.90)<sup>2</sup><br>80 | 0.61 (0.35; 0.83)<br>86 | 0.81 (0.45; 1.05)<sup>2</sup><br>88 |
| Euroimmun | 0.32 (0.65; 5.21)<br>65 | 4.60 (1.92; 7.25)<br>86 | 6.19 (2.45; 7.29)<br>88 |
| Roche | 2.94 (0.45; 12.27)<sup>3,4</sup><br>60<sup>§,&</sup> | 12.55 (3.02; 39.2)<sup>3</sup><br>88<sup>§</sup> | 34.17 (6.34; 43.26)<sup>4</sup><br>91<sup>&</sup> |
| Abbott | 3.02 (1.99; 5.21)<sup>5</sup><br>80 | 5.61 (2.01; 7.65)<sup>6</sup><br>81 | 6.82 (4.75; 7.98)<sup>5,6</sup><br>91 |
| DiaSorin | 37.00 (5.99; 67.80)<sup>7,8</sup><br>65 | 70.40 (26.5; 134)<sup>8</sup><br>83 | 86.1 (35.9; 154)<sup>8</sup><br>88 |
| LIPS S-RBD | 4.78 (2.14; 22.03)<br>75 | 14.19 (4.63; 58.10)<br>83 | 14.36 (6.03; 30.86)<br>91 |
| LIPS N | 5.20 (2.83; 10.49)<sup>9,10</sup><br>80 | 11.33 (6.24; 24.74)<sup>9,11</sup><br>95 | 19.49 (9.12; 80.20)<sup>10,11</sup><br>94 |
| Biosensor | NA<br>40<sup>£,€</sup> | NA<br>65<sup>£</sup> | NA<br>79<sup>€</sup> |

NA – not applicable
Statistical difference between quantitative data: <sup>1</sup>p=0.01; <sup>2</sup>p=0.026; <sup>3</sup>p=0.006; <sup>4</sup>p<0.001; <sup>5</sup>p=0.004; <sup>6</sup>p=0.04; <sup>7</sup>p=0.017; <sup>8</sup>p=0.008; <sup>9</sup>p=0.018; <sup>10</sup>p<0.001; <sup>11</sup>p=0.029, and between proportions of positive results: <sup>#</sup>p=0.01; <sup>□</sup>p=0.002; <sup>§</sup>p=0.02; <sup>&</sup>p=0.01; <sup>£</sup>p=0.002; <sup>€</sup>p=0.007

**Table 3.** Antibody detection by tests in COVID-19 patients (n=97) in different time to test groups

| Tests | Quantitative test value median (25%;75% percentiles)<br>% of positive tests in subgroup |  |  |
| --- | --- | --- | --- |
|  | 7-14 days<br>n=20 | 15-30 days<br>n=35 | 31-57 days<br>n=42 |
| SNIBE | 2.02 (0.23; 17.51)<br>55 | 4.21 (0.8; 17.94)<br>69 | 5.18 (1.19; 14.02)<br>79 |
| Epitope | 0.65 (0.3; 1.03)<br>80 | 0.73 (0.37; 0.96)<br>89 | 0.62 (0.36; 0.87)<br>86 |
| Euroimmun | 2.43 (0.68; 4.92)<sup>1,2</sup><br>55<sup>#,□</sup> | 5.18 (2.32; 7.88)<sup>1</sup><br>89<sup>#</sup> | 4.81 (2.40; 6.83)<sup>2</sup><br>90<sup>□</sup> |
| Roche | 3.18 (1.15; 8.24)<sup>3,4</sup><br>75 | 15.05 (6.02; 51.45)<sup>3</sup><br>83 | 31.90 (5.39; 43.16)<sup>4</sup><br>88 |
| Abbott | 4.27 (2.23; 6.77)<br>85 | 6.15 (2.67; 7.76)<br>86 | 5.88 (3.11; 7.65)<br>83 |
| DiaSorin | 27 (6.46; 73.55)<sup>5,6</sup><br>55<sup>§,&</sup> | 98.8 (28.7; 198)<sup>5</sup><br>86<sup>§</sup> | 70.15 (30.2; 134)<sup>6</sup><br>90<sup>&</sup> |
| LIPS S-RBD | 3.05 (1.85; 6.08)<sup>7,8</sup><br>70 | 22.69 (9.31; 59.60)<sup>7</sup><br>91 | 13.48 (4.68; 34.15)<sup>8</sup><br>86 |
| LIPS N | 10.49 (4.14; 35.73)<br>90 | 8.48 (3.38; 13.11)<sup>9</sup><br>89 | 16.13 (9.12; 63.90)<sup>9</sup><br>95 |
| Biosensor | NA<br>60 | NA<br>69 | NA<br>64 |

NA – not applicable
Statistical difference between quantitative data: <sup>1</sup>p=0.006; <sup>2</sup>p=0.01; <sup>3</sup>p=0.003; <sup>4</sup>p<0.001; <sup>5</sup>p=0.003; <sup>6</sup>p=0.01; <sup>7</sup>p<0.001; <sup>8</sup>p<0.001; <sup>9</sup>p=0.005, and between proportions of positive results: <sup>#</sup>p=0.008; <sup>□</sup>p=0.003; <sup>§</sup>p=0.02; <sup>&</sup>p=0.003

### Relations between antibody detection and time to test (TTT)

We found significant correlation between TTT and test quantitative result in case of Roche (ρ=0.38; p<0.001), Euroimmun (ρ=0.21; p=0.04) and LIPS N (ρ=0.28; p=0.009) test. We also described significant differences between TTT groups in quantitative test data as well as in positivity rate in case of some but not all tests (Table 4).

### Relations between antibody detection and patients’ age and sex

No any correlation between test results and patients’ age or sex were found.

## Discussion

We found that different tests gave diverse antibody results if applied on heterogeneous COVID-19 patient group. In less than 60% of COVID-19 cases all tests gave identical positive or negative result. If assumed that all COVID-19 patients (confirmed by positive SARS-CoV-2 PCR test) should develop IgG antibodies, the sensitivity of tests varied from 65 to 92% that is much less than reported by the manufacturers (1). However, similarly low sensitivity rates have been reported in recent meta-analysis (2). A combination of 2 tests with different protein markers, e.g. re-testing close to cut-off Abbott (N protein) IgG negative samples by DiaSorin (S1 and S2 proteins), as practiced in SYNLAB Estonia, somewhat added higher analytical sensitivity to COVID-19 diagnostics. Although this combination did not affect the specificity in our control group, comparisons in larger study groups should be done. However, even if several different tests are combined, some confirmed COVID-19 cases remain negative for antibodies.

Correlation and agreements between the tests studied here varied highly in our COVID-19 group. The best correlation was found between Epitope and DiaSorin, which could be explained by detecting the antibodies to the same viral antigen (spike protein), however the relationship was not entirely clear since probably different subunits of N and S proteins have been used in tests and no exact data are available from manufacturers. One of the weakest agreements in positive/negative results and absence of correlation in quantitative test values was found when comparing IgG antibodies to nucleocapsid and to RBD of spike protein detected by LIPS tests. It is thus plausible that the patients develop diverse IgG antibody reactivities to SARS-CoV-2 proteins and their epitopes.

While analyzing the results from different tests in patient groups with different symptoms scores we found that patients with more symptoms usually had higher positivity rate and higher levels of antibodies. This is in accordance with some previous studies (3). However, this relationship was not uniform in all tests. For example, LIPS detected significant differences in anti-N IgG levels between asymptomatic, pauci-symptomatic and polysymptomatic COVID-19 patients. At the same time anti-RBD IgG variation within the groups was higher and no significant differences between groups were found. Thus, it seems that the production of IgG antibodies against some virus proteins are more symptom-dependent than the others. The symptomatic patients may develop higher immune response (and probably protection) than asymptomatic, but at the same time diagnosing of late or previous of COVID-19 by antibody test may be problematic in asymptomatic and pauci-symptomatic patients due to low antibody level. For some tests (such as Abbott), the absence of clinical severity seems not to affect significantly positivity rate, but for others (such as Biosensor rapid test or SNIBE CLIA) positivity rate in asymptomatic COVID-19 cases was about two times lower than polysymptomatic ones. More studies are needed to confirm the finding that some antibody tests (that use specific antigens) are more suitable to diagnose asymptomatic COVID-19 cases than others.

We also found some test-dependent differences while comparing antibody detection among TTT groups. For example anti-N IgG was detected by Abbott in high level already at 7-14 days TTT group and no differences were found with later TTT groups, but in other tests (such as Euroimmun) lower detection rate and antibody levels were associated with shorter TTT. The reason could be in an analytical sensitivity of the test and also in usage of different viral proteins – antibodies to some virus proteins may appear earlier and at increased levels than others.

Our study has several limitations. Firstly, although the onset of disease could be dated relatively precisely in symptomatic cases, in asymptomatic cases the disease detection depends on random PCR screening of risk groups. Thus, the first positive SARS-CoV-2 PCR is not equivalent to disease onset and one should be careful drawing conclusions based of TTT in such heterogeneous group of patients that includes symptomatic and asymptomatic patients. Secondly, we only analyzed IgG and total antibody values. The main reasons were the absence of IgM tests from several manufacturers (Abbott, DiaSorin) in the time of testing and questionable reliability of available ones. SNIBE and Epitope IgM tests gave significantly lower positive results than IgG tests and added no additional positive cases (all IgM positive cases were also IgG positive). Euroimmun IgA had high positivity rate (ca 90% in COVID-19 patients) but also high proportion of nonspecific reactions (21% of positive and borderline cases) in pre COVID-19 control group. Thus, until reliable commercial IgM and IgA tests are available, we can’t evaluate the role of these antibodies in infection and applicability of these tests in clinical practice or surveillance studies.

In conclusion, our study gave new theoretical insight to COVID-19 diagnostic testing and has practical implications. We confirmed that SARS-CoV-2 antibody response depends on clinical symptoms and time of testing, but we also found that this relation is dependent on test setup and viral antigens used in the tests. This means that not all antibody tests work uniformly well in symptomatic and asymptomatic cases and in different time periods from disease onset. This explains some contradictory results of previous studies and should be taken into consideration in clinical practice and epidemiological studies.

## Data Availability

All data is available by request

## Acknowledgements

We thank Keith Orchard for language corrections.

## Ethical approval

Study has been approved by Ethical Committee of Tartu University on April 23, 2020 (nr 311/T-1).

## Contributors

PN and ES: general study design and writing the manuscript; KH, JH and IE: performance and interpretation of laboratory antibody tests, critical reading of manuscript; JP: organizing patients selection and sample collection, analyzing patients data; LH, PR and PP: development and performance of in-house test and critical reading of manuscript; PJ: preparation of study design and ethical aspects; KT: statistical analysis and writing the manuscript.

